# The genotype-phenotype correlation of NR5A1 variants in 46,XY individuals: a study protocol

**DOI:** 10.1101/2024.08.27.24312633

**Authors:** Renata Thomazini Dallago, Rafael Loch Batista, Berenice Bilharinho de Mendonça, Vania dos Santos Nunes Nogueira

## Abstract

**Introduction:** Disorders of sex development (DSDs) were defined as congenital conditions in which the development of chromosomal, gonadal and anatomic sex is atypical. Nuclear receptor subfamily 5 group A member 1 (*NR5A1*), previously known as steroidogenic factor 1 (*SF1*) plays a crucial role in transcriptional regulation of genes involved in steroidogenesis, adrenal and gonadal development, and reproduction. *NR5A1* emerged to be causative of 10 to 20% of 46,XY DSD.

**Objective:** The present study aims to perform a systematic review evaluating the association between phenotype and genotype in patients with *NR5A1* defects and 46,XY DSD looking for outcomes of spontaneous puberty, change of gender and social sex.

**Method:** The proposed systematic review will be conducted in accordance with the Joanna Briggs Institute methodology for systematic reviews of etiology and risk. This review will consider observational studies, including prospective and retrospective cohort studies, cross sectional, case series and case reports studies. We will focus on studies that included patients with allelic variants in the *NR5A1* gene and 46,XY DSD. This review will consider studies that include the outcomes of spontaneous puberty, gender change and genotype/phenotype correlation associating the types of variants with the degree of external genitalia virilization.

**Conclusion:** Results of this review can help in the management and clinical understanding of patients with *NR5A1* defects and 46,XY DSD.

## 1. Introduction

Disorders of sex development (DSDs) were defined as congenital conditions in which the development of chromosomal, gonadal and anatomic sex is atypical (1). DSD are often detected at birth with a newborn presenting with ambiguous genitalia (estimated incidence of 1 in 4,500-5,5000) or in adolescence with atypical pubertal development (2), 75% of whom have a 46,XY karyotype (3).

Nuclear receptor subfamily 5 group A member 1 (*NR5A1*), previously known as steroidogenic factor 1 (*SF1*), is a gene located on chromosome 9q33.3 which codifies the *SF1* protein (OMIM 184757) (4). The *NR5A1* plays a crucial role in transcriptional regulation of genes involved in steroidogenesis, adrenal and gonadal development, and reproduction (5)(6). *NR5A1* emerged to be causative of 10 to 20% of 46,XY DSD (4).

The first two mutations described in *NR5A1* gene were identified in 46,XY patients with primary adrenal insufficiency, phenotypically female with complete gonadal dysgenesis (CGD) and persistence of Müllerian ducts (7)(8). Meanwhile, numerous patients with genetic variations in the *NR5A1* gene have been described presenting with an extraordinary broad phenotypic spectra including DSD, male factor infertility, streak-like gonads, primary ovarian insufficiency and spleen anomalies (5)(9)(10)(11)(12). Although the crucial role of SF-1 in gonadal and adrenal development is well established, the broad clinical manifestation remains elusive with no correlation between phenotype and genotype (10)(13)(14).

To date, knowledge about gonadal function at puberty in patients with 46,XY DSD and *NR5A1* defects is very scarce, as most cases identified have undergone gonadectomy in early infancy (15). Finally, it’s common for the development of spontaneous signs of virilization at the age of puberty (15)(16). Testosterone concentrations differ between patients with *NR5A1* mutations as well, in most cases testosterone is low in the neonatal period but normal at puberty (4). Despite testosterone serum concentrations within the male reference range and spontaneous progression of puberty, the male patients frequently show impaired testicular growth. Therefore, testicular volume does not correlate well with the preserved Leydig cell function at puberty (16). Such as, despite normal testosterone levels, LH and FSH levels were elevated in most patients with further increase during the course of puberty (16). The almost normal testosterone levels after hCG stimulation or at pubertal age suggest that NR5A1 action may be less implicated in pubertal steroidogenesis than during fetal life (17).

Thus, the objective of the present study is to perform a systematic review evaluating the association between phenotype and genotype in patients with *NR5A1* defects and 46,XY DSD looking for outcomes of spontaneous puberty, change of gender and social sex.

## 2. Review question

The question that will be addressed in this review is what is the phenotypic implication of patients with an allelic variant in the *NR5A1* gene 46,XY DSD.

## 3. Methods

The proposed systematic review will be conducted in accordance with the Joanna Briggs Institute methodology for systematic reviews of etiology and risk (Chapter 7: Systematic reviews of etiology and risk) (18).

### 3.1. Inclusion criteria

#### Participants

This review will consider studies that included patients with allelic variants in the *NR5A1* gene and 46,XY DSD.

#### Exposure of interest

The exposure of interest is pathogenic or likely pathogenic allelic variant in the

*NR5A1* gene in 46,XY DSD patients.

#### Outcomes

This review will consider studies that include the following outcomes:

a. Spontaneous puberty The spontaneous puberty will be considered as a single measurement of testosterone level or luteinizing hormone with the following values in patients with puberty signs:
  a. Testosterone level above 150ng/dl
  b. Luteinizing hormone above 0,3U/L (ICMA) or 0,6U/L (IFMA)
b. Gender change The gender change will be considered when it occurs at the patient desire and not as a medical suggestion:
  a. Assigned sex
  b. Social sex
    i. Female
    ii. Male
c. Genotype/phenotype correlation associating the types of variants with the degree of external genitalia virilization:
  a. For the genitalia classification will be used the following criteria:
    i. Female: normal female genitalia
    ii. Atypical: with any sign of difference in the sex development like hypospadias, micropenis, clitoromegaly or partial labial fusion.
  b. Allelic variant type:
    i. Missense
    ii. Non-missense
      1. Nonsense
      2. Splicing
      3. Small indels
      4. CNV (Copy number variation).

### 3.2 Types of studies

This review will consider observational studies, including prospective and retrospective cohort studies, cross sectional, case series and case reports studies.

### 3.3. Exclusion criteria

This review will not consider patients with 46,XX DSD, sex chromosome DSD, allelic variant VUS, likely benign or benign.

Abstract of congress will be excluded.

### 3.4. Search strategy

The search strategy aims to identify published and unpublished studies. A preliminary search of PubMed was performed to identify articles on this topic. The search strategy, including all the identified keywords and index terms, will be adapted for each included information source. The reference lists of all studies selected for critical appraisal will be screened for additional eligible studies. There will be no language or year restrictions. There will be a search in national and international theses and dissertations.

#### 3.4.1. Information sources

Search strategies have been applied to the following electronic health databases: Embase (by Elsevier, 1980–2024), Medline (by PubMed, 1966–2024), HGMD (Human Gene Mutation Database) and I-DSD registry. We have used the following index terms and synonyms: disorders of sex development, disorder of sex development 46,XY, gonadal dysgenesis 46,XY, Steroidogenic factor 1, SF1 protein, human, NR5A1 protein, human. The draft PubMed and Embase search strategies are included in Appendix I.

#### 3.4.2. Study selection

All identified citations will be collated and uploaded into the bibliographic software Endnote 2020 and duplicates will be removed. Titles and abstracts will then be screened by two independent reviewers (RTD and RLB) using the free web application Rayyan QCRI. The full texts of the selected citations will be assessed in detail against the inclusion criteria by two independent reviewers. The reasons for the exclusion of full text studies will be recorded and reported in the systematic review. Disagreements between reviewers at each stage of the study selection process will be resolved through discussion or by a third reviewer (BBM). The results of the search will be reported in full in the final systematic review and presented in a Preferred Reporting Items for Systematic Reviews and Meta-analyses (PRISMA) flow diagram.

#### 3.4.3. Assessment of methodological quality

Eligible studies will be critically appraised by two independent reviewers (RTD and RLB) at the study level or methodological quality in the review, using standardized critical appraisal instruments from the Joanna Briggs Institute for cohort, cross sectional, and case series studies. Authors of papers will be contacted to request missing or additional data for clarification where required. Any disagreements that arise will be resolved through discussion or by a third reviewer. The results of the critical appraisal will be reported in narrative form and in a table. All studies, regardless of their methodological quality, will undergo data extraction and synthesis (where possible). If possible, the results of critical appraisal will be incorporated into sensibility analysis using a meta-analysis approach.

#### 3.4.4. Data extraction

Data will be extracted from the papers included in the review using a standardized data extraction tool by two independent reviewers (RTD and RLB). The extracted data will include specific details about exposure (age of hormone measures, age of clinical evaluation, age of gender change), study design, number of patients, number of pregnant women and outcome results.

#### 3.4.5. Data synthesis

Similar outcomes in at least two studies will be plotted in the meta-analysis using Stata Statistical Software 18 (Stata Statistical Software: Release 18. College Station, TX, USA). In controlled studies, for dichotomous data, relative risk will be calculated with 95% confidence intervals (CIs) as an estimate of the exposure effect. Continuous data will be expressed as means and standard deviations, and the differences between the means with 95% CIs will be used as an estimate of the exposure effect. A random effect model will be used for the meta-analysis. Narrative synthesis will be provided when quantitative synthesis is not appropriate.

Inconsistencies between the results of the studies included will be ascertained by applying the chi-square test (c2). For c2, statistical heterogeneity will be considered if p <0.10.

## 4. Conclusion

Results of this review can help in the management and clinical understanding of patients with *NR5A1* defects and 46,XY DSD.

## Data Availability

All data produced in the present work are contained in the manuscript

## Ethical approval

As no primary data collection will be undertaken, no formal ethical assessment is required by the authors institution.

## Consent

This study does not involve human participants

## Funding

This research has been partially supported by the São Paulo Research Foundation (FAPESP) (grant number 19/26780-9).

## Acknowledgements

This protocol was performed as a result of the Comprehensive Systematic Review Training Program (CSRT), ministered by the Brazilian Center for Evidence-based Healthcare: A JBI Center of Excellence.

## Competing interests

Authors have declared that no competing interests exist.

## Appendix

**Appendix I: Search strategy**

**Pubmed =** “Disorders of Sex Development”[Mesh] OR (Sex Development Disorders) OR (Sex Development Disorder) OR (Disorders of Sexual Development) OR (Sexual Development Disorders) OR (Sexual Development Disorder) OR (Ambiguous Genitalia) OR (Genitalia, Ambiguous) OR (Genital Ambiguity) OR (Ambiguities, Genital) OR (Ambiguity, Genital) OR (Genital Ambiguities) OR (Intersex Conditions) OR (Condition, Intersex) OR (Conditions, Intersex) OR (Intersex Condition) OR (Pseudohermaphroditism) OR (Sex Differentiation Disorders) OR (Differentiation Disorder, Sex) OR (Differentiation Disorders, Sex) OR (Disorder, Sex Differentiation) OR (Disorders, Sex Differentiation) OR (Sex Differentiation Disorder) OR (Sexual Differentiation Disorders) OR (Differentiation Disorder, Sexual) OR (Differentiation Disorders, Sexual) OR (Disorder, Sex Differentiation) OR (Disorders, Sex Differentiation) OR (Sexual Differentiation Disorder) OR (Hermaphroditism) OR (Intersexuality) OR (Intersexualities) OR “Disorder of Sex Development, 46,XY”[Mesh] OR (46,XY Disorders of Sex Development) OR (46,XY DSD) OR (46,XY DSDs) OR (DSD, 46,XY) OR (DSDs, 46,XY) OR (46, XY Disorders of Sex Development) OR (46, XY DSD) OR (46,XY Sex Reversal 3) OR (46,XY Sex Reversal, Partial or Complete,

NR5A1-Related) OR (Sex Reversal, XY, With Or Without Adrenal Failure) OR (46,XY Gonadal Dysgenesis, Complete or Partial, With or Without Adrenal Failure) OR (Male Pseudohermaphroditism) OR (Male Pseudohermaphroditisms) OR (Pseudohermaphroditism, Male) OR (Pseudohermaphroditisms, Male) OR “Gonadal Dysgenesis, 46,XY”[Mesh] OR (Sex Reversal, Gonadal, 46, XY) OR (Gonadal Dysgenesis, 46, XY) OR (46, XY Gonadal Dysgenesis) OR (46, XY Gonadal Sex Reversal) OR (Swyer Syndrome) OR (Syndrome, Swyer) OR (Pure Gonadal Dysgenesis 46,XY) OR (46,XY Complete Gonadal Dysgenesis) OR (XY Pure Gonadal Dysgenesis) OR (Complete Gonadal Dysgenesis, 46, XY) OR (Pure Gonadal Dysgenesis, 46, XY) AND “Steroidogenic Factor 1”[Mesh] OR (Ad4-Binding Protein) OR (Ad4 Binding Protein) OR (Adrenal 4 Binding Protein) OR (AD4BP Protein) OR (Fushi Tarazu Factor Homolog 1) OR (NR5A1 Protein) OR (Nuclear Receptor 5A1 Protein) OR (FTZF1 Protein) OR (SF-1 Transcription Factor) OR (Transcription Factor, SF-1) OR (Steroid Hormone Receptor Ad4BP) OR “SF1 protein, human” [Supplementary Concept] OR (splicing factor 1 protein, human) OR (zinc finger protein 162, human) OR (ZFM1 protein, human) OR (ZNF162 protein, human) OR “NR5A1 protein, human” [Supplementary Concept] OR (FTZF1 protein, human) OR (steroidogenic factor 1, human) OR (nuclear receptor subfamily 5, group A, member 1 protein, human) OR (AD4BP protein, human) OR (fushi tarazu factor (Drosophila) homolog 1 protein, human) = 2,220 results.

**Embase =** ‘disorder of sex development’/exp OR ‘disorder of sex development’/exp OR ‘46, XX disorders of sex development’ OR ‘46, XX testicular disorders of sex development’ OR ‘46, XY disorders of sex development’ OR ‘46, XX DSD’ OR ‘46, XY disorder of sex development’ OR ‘46, XY DSD’ OR ‘difference of sex development’ OR ‘differences of sex development’ OR ‘disorder of sex development, 46, XY’ OR ‘disorder of sex differentiation’ OR ‘disorder of sexual development’ OR ‘disorder of sexual differentiation’ OR ‘disorders of sex development’ OR ‘disorders of sex differentiation’ OR ‘disorders of sexual development’ OR ‘disorders of sexual differentiation’ OR ‘divergence of sex development’ OR ‘divergences of sex development’ OR ‘DSD (disorder of sex development)’ OR ‘intersexualism’ OR ‘intersexuality’ OR ‘ovotesticular disorders of sex development’ OR ‘sex chromosome disorder of sex development’ OR ‘sex chromosome disorders of sex development’ OR ‘sex development disorder’ OR ‘sex development disorders’ OR ‘sex differentiation disorder’ OR ‘sex differentiation disorders’ OR ‘sex differentiation disturbance’ OR ‘sexual development disorder’ OR ‘sexual development disorders’ OR ‘sexual differentiation disorder’ OR ‘sexual differentiation disorders’ OR ‘disorder of sex development’ OR ‘XY gonadal dysgenesis’/exp OR ‘46, XY CGD’ OR ‘46, XY complete gonadal dysgenesis’ OR ‘46, XY gonadal dysgenesis’ OR ‘46, XY pure gonadal dysgenesis’ OR ‘complete gonadal dysgenesis, 46, XY’ OR ‘gonadal dysgenesis, 46, XY’ OR ‘gonadal dysgenesis, pure 46xy’ OR ‘pure gonadal dysgenesis 46, XY’ OR ‘Swyer syndrome’ OR ‘Swyer’s syndrome’ OR ‘XY pure gonadal dysgenesis’ OR ‘XY gonadal dysgenesis’ AND ‘steroidogenic factor 1’/exp OR ‘adrenal 4 binding protein’ OR ‘steroidogenic factor 1’ OR ‘NR5A1 protein’/exp OR ‘nr5a1 gene’ OR ‘nr5a1 protein human’ OR ‘ftz f1 gene’ OR ‘sf 1 gene’= 428 results

## Notes

### Competing Interest Statement

The authors have declared no competing interest.

### Funding Statement

This study did not receive any funding

